# Activation of the renin-angiotensin-aldosterone system is attenuated in hypertensive compared to normotensive pregnancy

**DOI:** 10.1101/2022.11.26.22282783

**Authors:** Robin Shoemaker, Marko Poglitsch, Hong Huang, Katherine Vignes, Aarthi Srinivasan, Cynthia Cockerham, Aric Schadler, John A. Bauer, John M. O’Brien

**Affiliations:** Department of Dietetics and Human Nutrition, University of Kentucky, Lexington KY, USA; Attoquant Diagnostics GmbH, Vienna, Austria; Department of Pediatrics, University of Kentucky, Lexington KY, USA; Division of Maternal Fetal Medicine, Department of Obstetrics and Gynecology, University of Kentucky, Lexington KY, USA

**Keywords:** angiotensin, maternal, pregnancy hypertension, biomarkers, mass spectrometry, aldosterone

## Abstract

Hypertension during pregnancy increases the risk for adverse maternal and fetal outcomes, but mechanisms of pregnancy hypertension are not precisely understood. Elevated plasma renin activity and aldosterone concentrations play an important role in the normal physiologic adaptation to pregnancy. These effectors are reduced in patients with pregnancy hypertension, creating an opportunity to define features of the renin-angiotensin-aldosterone system (RAAS) that are characteristic of this disorder. In the current study, we used a novel LC-MS/MS-based methodology to develop comprehensive profiles of RAAS peptides and effectors over gestation in a cohort of n=74 pregnant women followed prospectively for the development of gestational hypertension and pre-eclampsia (HYP, n=27) versus remaining normotensive (NT, n=47). In NT pregnancy, the plasma renin activity surrogate, (PRA-S, calculated from the sum of [angiotensin I] + [angiotensin II) and aldosterone concentrations significantly increased from first to third trimester, accompanied by a modest increase in concentrations of angiotensin peptide metabolites. In contrast, in HYP pregnancies PRA-S and angiotensin peptides were largely unchanged over gestation, and third trimester aldosterone concentrations were significantly lower compared to NT pregnancies. Results indicate that the predominate features of pregnancies that develop HYP are stalled or waning activation of the RAAS in the second half of pregnancy (accompanied by unchanging levels of angiotensin peptides) and attenuated secretion of aldosterone.

## 1. Introduction

New onset hypertension during pregnancy (gestational hypertension or pre-eclampsia) affects 5-10% of pregnancies, and the prevalence in the United States is rising[1]. Development of hypertension during pregnancy is associated with long-term health risks for both the mother and the offspring[2], and increases maternal risk for cardiovascular diseases later in life[3, 4]. Knowledge of mechanisms underlying perinatal hypertension could lead to improved strategies for the treatment and/or management of hypertension during pregnancy [5, 6].

The renin angiotensin aldosterone system (RAAS) is a master regulator of blood pressure and fluid homeostasis in pregnant and non-pregnant states[7, 8]. Activity of the RAAS is initiated by the enzymatic cleavage of a ten-amino acid peptide, angiotensin I (Ang I) from the N-terminus of angiotensinogen (AGT) by the enzyme renin in a highly regulated reaction. The rate of Ang I formation in plasma, also called plasma renin activity (PRA), is determined by the concentrations of active renin and AGT, where renin is rate-limiting. Ang I serves as a substrate for angiotensin-converting enzyme (ACE), resulting in the formation of angiotensin II (Ang II), the primary effector molecule of the RAAS. The concentration of active Ang II is controlled via negative feedback on renin [9], and by metabolism at the n- or c-terminal by amino- and carboxy peptidases in plasma, where the latter is considered to be a counter-regulatory arm of the RAAS. Cleavage of Ang II by angiotensin-converting enzyme 2 (ACE2) generates angiotensin-1-7(Ang 1-7)[10], a peptide with purported vasodilatory and other counter-effects to Ang II[11].

The classical effects of Ang II are mediated through the Ang II type 1 (AT_1_) receptor, and include vasoconstriction, activation of the sympathetic nervous system, and stimulation of aldosterone release from the adrenal cortex[12, 13]. Outside of pregnancy, excess effects of Ang II and aldosterone are associated with inflammation, fibrosis, and endothelial dysfunction[14, 15], and abnormal function of the counter-regulatory arm of the RAAS has been described in many cardiovascular [16, 17], renal[18], and infectious disease states(i.e. COVID-19)[19].

In healthy pregnancy, there is pronounced activation of the RAAS, where PRA and aldosterone concentrations are increased compared to non-pregnant states[20–22]. Upregulation of renin activity and aldosterone secretion play key roles in physiologic adaptations to healthy pregnancy, such as promoting water and sodium reabsorption sufficient to support the demand for a 40-50% expansion in blood volume[8, 23, 24]. In contrast, numerous studies have demonstrated that pregnancies with pre-eclampsia or gestational hypertension have lower plasma renin activity and lower aldosterone concentrations compared to normotensive pregnancies. [21, 25–27]. Limited studies suggest the alternative RAAS may also play a role in pregnancy, and that alterations in these components, e.g. Ang-1-7 and/or ACE2, are associated with pre-eclampsia and small-for-gestational age infants[28–30]. The course of RAAS activity over the course of gestation is not precisely understood in either normal or hypertensive pregnancy[23, 31]. Understanding key deviations in the normal activity of the RAAS in pregnancy could lead to improved detection or management of gestational hypertension and/or pre-eclampsia.

RAS Fingerprint^TM^ is novel, liquid chromatography-tandem mass spectrometry (LC-MS/MS)-based methodology for simultaneous quantification of multiple components of the RAAS in a single sample. Outside of pregnancy, recent studies have utilized RAS Fingerprint^TM^ (or RAS profiling) to generate a biochemical “snapshot” of RAAS activity reflective of disease state in patients with hypertension[32], heart failure[33, 34], and other diseases[35, 36]. In the current study, we used RAS Fingerprint to quantify key effectors (angiotensin peptides and aldosterone) of the RAAS in a cohort of women followed prospectively for the development of gestational hypertension and pre-eclampsia. Our objective was to define prominent features of RAAS activity associated with the development of pregnancy hypertension.

## 2. Methods and Materials

### Study Population

This is a secondary analysis of n=74 pregnant individuals enrolled in an ongoing study at the University of Kentucky. All subjects gave informed consent to participate in the study, which was approved by the University of Kentucky Institutional Review Board, and all study procedures/methods were performed in accordance with relevant guidelines and regulations. The cohort is comprised of pregnant individuals without pre-pregnancy hypertension, but otherwise meeting the following clinical risk factors for pre-eclampsia[37], where inclusion in the study were more than one of the following clinical risk factors: history of preterm preeclampsia, type 1 or 2 diabetes, renal or autoimmune disease; or more than two of the following clinical risk factors: nulliparity, obesity (BMI > 30), family history of pre-eclampsia, Black race, lower income, age 35 or older, previous pregnancy with small birth weight or adverse outcome, or in-vitro conception. Patients not meeting these criteria were recruited as controls. Exclusion criteria included age less than 18 or greater than 45 years, multifetal gestation, history of allergy to aspirin, gastrointestinal bleeding, severe peptic ulcer or liver dysfunction, patients on anticoagulant medications, and women with anomalous fetus.

### Study Design and Groups

Pregnant women between 18 and 45 years of age in the first trimester of pregnancy were recruited at the time of their routine prenatal visits or ultrasound appointments in the first trimester screening and followed prospectively for the development of pre-eclampsia, gestational hypertension, and related pregnancy outcomes. Patients were treated according to standard clinical guidelines and data collected as part of routine clinical care. Demographic information was collected upon enrollment. Clinical data and maternal blood were collected at routine visits by trained study personnel at the first (11-16 weeks) and third (28-32 weeks) trimesters of pregnancy, and outcomes were recorded at delivery. Blood pressure was measured following the American Heart Association Guidelines[38], and was determined as the average of two consecutive measurements per arm assessed in a seated, resting position by one observer. Clinical data and outcomes were obtained from electronic medical records (EMR), and input into a REDcap database by study personnel. Maternal blood was collected into red-top serum tubes and rested at room temperature for 60 minutes followed by centrifugation, transfer into 200 uL single-use aliquots, and barcoding. Samples were stored at - 80°C until analysis.

Development of pre-eclampsia or gestational hypertension was the primary endpoint. Pregnancy outcomes were determined after delivery by trained clinical staff. Pre-eclampsia was diagnosed according to guidelines by the American College of Gynecologists[39]: either systolic blood pressure (SBP) or diastolic blood pressure (DBP) greater than 140 or 90 mmHg, respectively, measured at least four hours apart after 20 weeks gestation, and one of the following: proteinuria of 300 mg in 24 hours or a urine to protein creatinine ratio = 0.3, elevated serum creatine greater than 1.1 mg/dL, liver function tests (AST/ALT) double the upper limits of normal, platelet counts of <100,00/microliter, flash pulmonary edema, or neurologic features including blurry vision or headaches not relieved with medication. Gestational hypertension was diagnosed as either SBP or DBP greater than 140 or 90 mmHg, respectively, after 20 weeks gestation in women with previously normal blood pressure[39]. The following adverse outcomes were also recorded: gestational diabetes, small for gestational age (defined as fetal weight below the 10^th^ percentile), postpartum hemorrhage, preterm premature rupture of membranes (PPROM), pre-term labor or pre-term delivery (<34 weeks), presence of polyhydramnios, or fetal death.

In the current study, patients were grouped for analysis based on whether they met the primary endpoint of either pre-eclampsia or gestational hypertension (HYP, n=27), or remained normotensive (NT, n=47). We then investigated differences in measured serum components of the RAAS over gestation (in the first versus third trimester) and at each time point among NT and HYP groups.

### Quantification of angiotensin peptides, aldosterone, and equilibrium-based biomarkers PRA-S, ACE-S, and the aldosterone to Ang II ratio (AA2-R) in serum using RAS Fingerprint^TM^

Serum concentrations of six angiotensin peptides (Ang I, Ang II, Ang III, Ang IV, Ang-1-5, and Ang-1-7) and aldosterone were quantified by LC-MS/MS following a controlled ex vivo equilibration (i.e. equilibrium analysis) that has been previously described[40–43]. Equilibrium analysis generates a “snapshot” of RAAS activity in a sample that reflects the biochemical features of the circulating RAAS, based on the principle that all components required to generate angiotensin metabolites in vivo are present in plasma. Briefly, serum samples were incubated at 37 °C for one hour to generate a controlled equilibrium, followed by stabilization through addition of an enzyme inhibitor cocktail. Samples were spiked with stable isotope-labeled internal standards at 200 pg/mL for each angiotensin metabolite. The samples were subjected to C-18-based solid-phase-extraction, followed by LC-MS/MS analysis using a reversed-phase analytical column (Acquity UPLC C18, Waters, Milford, Massachusetts, USA) operating in line with a Xevo TQ-S triple quadruple mass spectrometer in multiple reaction monitoring mode (Waters, Milford, Massachusetts, USA). Internal standards were used to correct for peptide recovery of the sample preparation procedure for each analyte in each individual sample. Analyte concentrations were determined using software (MassLynx/Target/Lynx, Waters, Milford, Massachusetts, USA) via integration of the total ion chromatogram obtained from a sum of quantifier transitions that have been optimized for sensitivity and specificity, and integrated signals exceeded a signal-to noise ratio of 10. Analyte concentrations were determined via linear calibration and reported in pmol/L. Batch performance is evaluated based on calculated levels of calibrator and quality control samples (must be within 15% of nominal concentration); lowest calibrator (lower limit of quantification – LLOQ) must be within 20% of nominal concentration; at least 67% of all quality control and calibrators must comply. The lower limits of quantification (LLOQs) for each peptide were: angiotensin I: 5 pmol/L; angiotensin II: 4 pmol/L; angiotensin III: 4 pmol/L; angiotensin IV: 2 pmol/L; angiotensin-(1-5): 2 pmol/L; angiotensin-(1-7): 3 pmol/L, and aldosterone: 13 pmol/L. For values below the LLOQ, means for each analyte and calculation of equilibrium-based biomarkers were calculated using half the LLOQ[43].

Equilibrium concentrations of Ang I, Ang II, and aldosterone were used to calculate biomarkers reflecting PRA, ACE activity, and the aldosterone to Ang II ratio (AA2-R), as previously described[32, 41]. The plasma renin activity surrogate (PRA-S) was calculated from the sum of the equilibrium concentrations of Ang I and Ang II ([eqAng I] + [eqAng I] (pmol/L); the surrogate for ACE activity (ACE-S) was calculated from the ratio of the equilibrium concentrations of Ang II to Ang I ([eqAng II] / [eqAng I] (pmol/L/pmol/L); the aldosterone to Ang II ratio (AA2-R) was calculated by diving aldosterone concentrations by equilibrium concentrations of Ang II ([Aldosterone] / [eqAng II] (pmol/L/pmol/L). PRA-S, ACE-S, and AA2-R have been utilized as biomarkers of the RAAS that correlate with clinical parameters and/or predict outcomes in numerous studies, including in humans with hypertension and/or primary aldosteronism[32, 44–46], heart failure[33, 43, 47], COVID-19[48, 49], and others[50, 51].

### Statistical Analysis

Statistical analyses were performed via GraphPad Prism version 9.5.1 (San Diego, CA, USA) and SPSS Statistics version 24 (IBM, Armonk, NY). All data were assessed for normality, and the appropriate parametric/nonparametric tests used. Clinical and demographic data were analyzed for between-group differences (NT versus HYP), where continuous variables were analyzed using paired t-tests, and categorical variables were analyzed using chi-square or Fisher’s exact test. For serum components of the RAAS, Mann-Whitney tests were applied for between-groups analysis at each time point in independent samples, and within-group changes from first to third trimester were determined using Wilcoxon matched-pairs signed rank tests. A p-value less than 0.05 was considered statistically significant.

## 3. Results

### Subject characteristics

Paired clinical data and biological samples from first and third trimester visits were obtained for 74 women between April 2019 and May 2020. The cohort demographics, clinical characteristics, and outcomes are described in Table 1. The cohort was primarily Caucasian, with Black, Hispanic, and mixed-race individuals distributed fairly equally among groups. The HYP group had a slightly greater risk profile compared to NT, with more patients being primiparous, having a previous pre-eclamptic pregnancy, having type 1 diabetes, and taking aspirin for the prevention of pre-eclampsia. The NT group contained slightly more patients with type 2 diabetes, and with renal or autoimmune disease. First trimester BMI was significantly greater in HYP compared to the NT, as was first and third trimester SBP and DBP, although only four patients in the HYP received hypertension treatment. Duration of gestational was significantly shorter in HYP versus NT groups.

**Table 1.** Patient demographics and clinical characteristics of women that developed gestational hypertension or pre-eclampsia (HYP) in pregnancy or remained normotensive (NT).

| Parameter | NT<br>N=47 | HYP<br>N=27 |
| --- | --- | --- |
| **Patient Demographics, n (%)** |  |  |
| Race |  |  |
| Caucasian | 33 (70) | 20 (74) |
| Black or African American | 7 (15) | 4 (15) |
| Hispanic | 6 (13) | 2 (7) |
| Asian | 1 (2) | 0 |
| Mixed race | 0 | 1 (4) |
| Primiparous | 19 (40) | 14 (52) |
| Previous pre-eclampsia | 6 (13) | 8 (30) |
| Type 1 DM | 7 (15) | 9 (33) |
| Type 2 DM | 5 (11) | 2 (7) |
| Renal disease | 1 (2) | 0 |
| Autoimmune disease | 7 (15) | 0 |
| Medications |  |  |
| Aspirin | 22 (47) | 18 (67) |
| Labetalol or Nifedipine | 0 | 4 (15) |
| **Clinical Characteristics, mean ± SD** |  |  |
| Age, years | 30 ± 6 | 28 ± 6 |
| BMI (1<sup>st</sup> trimester), kg/m<sup>2</sup> | 29 ± 7 | 32 ± 8<sup>#</sup> |
| SBP, mmHg |  |  |
| 1<sup>st</sup> Trimester | 113 ± 11 | 122 ± 9<sup>###</sup> |
| 3<sup>rd</sup> Trimester | 115 ± 9 | 126 ± 10<sup>####</sup> |
| DBP, mmHg |  |  |
| 1<sup>st</sup> Trimester | 73 ± 8 | 80 ± 6<sup>###</sup> |
| 3<sup>rd</sup> Trimester | 73 ± 6 | 81 ± 6<sup>####</sup> |
| Gestational age at delivery, weeks | 38.6 ± 1.5 | 36.8 ± 1.6<sup>####</sup> |
| **Outcomes, n (%)** |  |  |
| Gestational hypertension | 0 | 23 (85) |
| Pre-eclampsia | 0 | 6 (22) |
| Gestational diabetes | 3 (6) | 4 (15) |
| PPROM | 3 (6) | 1 (4) |
| Polyhydramnios | 4 (9) | 2 (7) |
| IUGR | 2 (5) | 0 |
| Pre-term labor or delivery | 3 (6) | 0 |
| Postpartum hemorrhage | 7 (15) | 4 (15) |

The HYP group was comprised of twenty-three patients who developed gestational hypertension and six patients that developed pre-eclampsia. Both NT and HYP patients experienced adverse outcomes such as gestational diabetes, PPROM, polyhydramnios, and postpartum hemorrhage. Slightly more HYP than NT patients developed gestational diabetes, but no HYP patients had IUGR or pre-term labor/delivery, compared to two and three, respectively, in the NT group.

### Attenuated activation of the classical RAAS over gestation in pregnancies that develop hypertension versus remaining normotensive

We used RAS Fingerprint with equilibrium analysis to quantify angiotensin peptides, aldosterone concentrations, and biomarkers in serum of pregnant women in the first and third trimester of pregnancy. Table 2 list the median and IQR for all measured and calculated components of the RAAS in the cohort - in general, most components of the RAAS were significantly increased over gestation in NT, but not HYP, and tended to be lower in HYP versus NT in the third trimester.

**Table 2.** Serum concentrations of RAAS components in normotensive (NT) and hypertensive (HYP) groups in the first and third trimesters of pregnancy.

| RAAS Component | NT<br>N=47 | HYP<br>N=20 | P value<sup>®</sup> |
| --- | --- | --- | --- |
| Ang I, pmol/L<br>1<sup>st</sup> Trimester<br>3<sup>rd</sup> Trimester | 72.7 (45.9 – 112.1)<br>115.6 (69.2 – 172.7)<sup>****</sup> | 61.6 (36.2 – 104.1)<br>71.6 (51.2 – 145.1) | 0.645<br>0.080 |
| Ang II, pmol/L<br>1<sup>st</sup> Trimester<br>3<sup>rd</sup> Trimester | 165.3 (110.5 – 197.0)<br>180.9 (130.8 – 290.2)<sup>*</sup> | 143.9 (76.3 – 211.7)<br>141.9 (87.8 – 244.5) | 0.404<br>0.053 |
| Ang III, pmol/L<br>1<sup>st</sup> Trimester<br>3<sup>rd</sup> Trimester | 4.3 (2.0 – 9.2)<br>5.9 (2.0 – 10.1) | 2.0 (2.0 – 7.3)<br>2.0 (2.0 – 4.9) | 0.602<br>0.012 |
| Ang IV, pmol/L<br>1<sup>st</sup> Trimester<br>3<sup>rd</sup> Trimester | 8.6 (5.3 – 12.9)<br>10.1 (5.2 – 14.0) | 5.8 (3.4 – 10.1)<br>7.4 (4.6 – 11.9) | 0.107<br>0.149 |
| Ang-1-5, pmol/L<br>1<sup>st</sup> Trimester<br>3<sup>rd</sup> Trimester | 5.3 (3.4 – 7.8)<br>7.7 (5.3 – 10.9)<sup>**</sup> | 4.3 (3.0 – 8.3)<br>5.2 (3.3 – 8.2) | 0.571<br>0.084 |
| Aldosterone, pmol/L<br>1<sup>st</sup> Trimester<br>3<sup>rd</sup> Trimester | 386.1 (226.2 – 626.2)<br>970.5 (468.3 – 1586.0)<sup>****</sup> | 290.2 (181.3 – 791.0)<br>492.5 (260.3 – 826.9) | 0.680<br>0.003 |
| PRA-S, pmol/L<br>1<sup>st</sup> Trimester<br>3<sup>rd</sup> Trimester | 243.2 (157.5 – 309.1)<br>307.9 (214.7 – 474.8)<sup>***</sup> | 194.3 (126.8 – 341.7)<br>223.7 (143.4 – 343.2) | 0.426<br>0.067 |
| ACE-S, pmol/L / pmol/L<br>1<sup>st</sup> Trimester<br>3<sup>rd</sup> Trimester | 2.16 (1.83 – 2.98)<br>1.78 (1.30 – 2.08)<sup>****</sup> | 2.27 (1.79 – 3.21)<br>1.83 (1.45 – 2.41)<sup>**</sup> | 0.703<br>0.534 |
| Aldosterone-Ang II ratio<br>(AA2-R), pmol/L / pmol/L<br>1<sup>st</sup> Trimester<br>3<sup>rd</sup> Trimester | 2.07 (1.63 – 3.33)<br>4.25 (3.20 – 7.71)<sup>****</sup> | 2.25 (0.90 – 4.67)<br>3.38 (2.04 – 6.27) | 0.929<br>0.161 |

As previous studies of pregnancy indicated that classical biomarkers for activation of the RAAS, PRA (or direct renin concentration) and plasma aldosterone concentrations (PAC), are reduced in gestational hypertension or pre-eclampsia[25, 27, 52], we examined PRA-S and aldosterone concentrations generated using RAS Fingerprint in our cohort of NT and HYP pregnancies (Figure 1). PRA-S was significantly increased from the first to third trimester in pregnancies that remained NT (P<0.001) with a median percent increase of 31% (Figure 1A, 1C). In contrast, PRA-S was not statistically increased in pregnancies that developed HYP (p=0.889), and the median percent change in PRA-S over gestation in the HYP group was zero (Figure 1A, 1C). Median values for PRA-S were not different between NT and HYP at either the first or third trimester, although there was a trend toward a lower PRA-S in HYP versus NT in the third trimester (p=0.067; Figure 1A, Table 2). Similarly, serum aldosterone concentrations significantly increased over gestation in the NT group (P<0.0001), with a median percent increase of 135%. In HYP, aldosterone rose by 57%, although the change over gestation was not significant (p=0.141), and third trimester concentrations of aldosterone were significantly lower in HYP versus NT (P<0.01; Figure 1B, 1D, Table 2).

**Figure 1.**
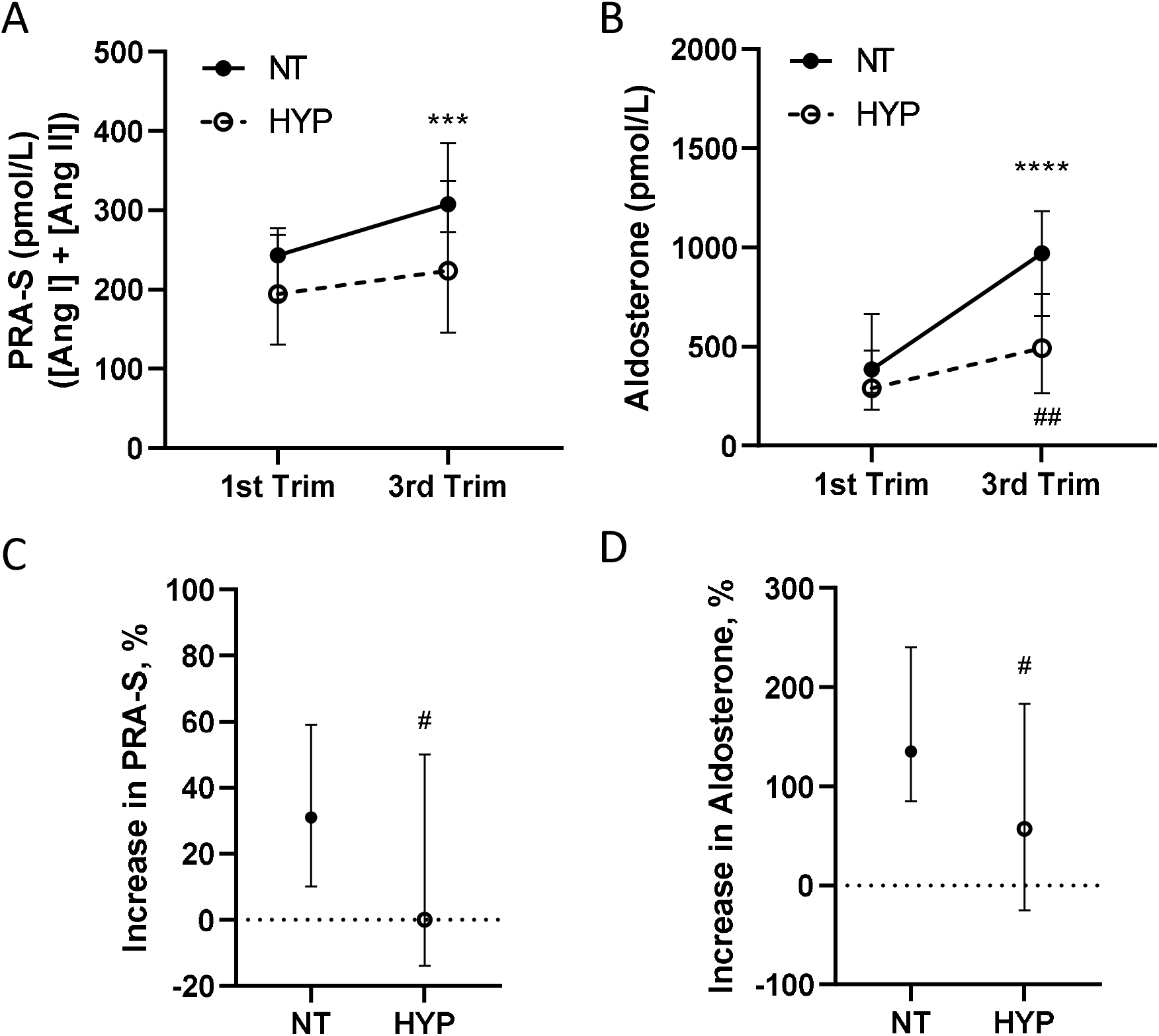
RAS Fingerprint-generated biomarkers for activation of the classical RAAS in normotensive versus hypertensive pregnancy. A) PRA-S, a surrogate for plasma activity calculated from the sum of the equilibrium concentrations of Ang I and Ang II, and B) aldosterone concentrations in the first and third trimesters of pregnancies that developed hypertension after 20 weeks gestation (HYP, n=27) and those that remained normotensive (NT, n=47). PRA-S and aldosterone levels were not different among NT and HYP in the first trimester, but increased over gestation only in the group that remained NT. Data are median, 95th confidence intervals (CI). ***, P<0.001, and ****, P<0.0001 for within-group increase from first to third trimester analyzed by Wilcoxon matched-pairs signed rank tests. ##, P<0.01 compared to NT using Mann-Whitney test. The percent change from first to third trimester in C)PRA-S and D) aldosterone concentrations in NT and HYP patients. The percent increase over gestation in PRA-S and aldosterone was significantly lower in pregnancies that developed HYP compared to NT. Data are median, 95th CI of the percent change for each biomarker over pregnancy in NT and HYP groups. #, P<0.05 compared to NT using Mann-Whitney test.

### Metabolism of Ang II and activation of the counter-regulatory RAAS in NT and HYP pregnancies

We quantified angiotensin metabolites upstream (Ang I) and downstream (Ang III and Ang IV, generated via n-terminal metabolism; and Ang-1-5 and Ang-1-7, generated via c-terminal metabolism) from Ang II (Figure 2A-D, Table 2). In NT, Ang II concentrations were only modestly increased across gestation (P<0.05), although Ang I formation was robustly increased across pregnancy (P<0.0001; Figure 2A, 2B, Table 2). In the HYP group, Ang II concentrations were not increased at all (p=0.594), but concentrations of Ang I were (non-significantly) increased over gestation (p=0.052; Figure 2A, 2B, Table 2). There was a relative decrease in Ang II to Ang I over gestation, reflected by a decrease in ACE-S, evident in both NT and HYP groups (Table 2). There was a relative increase in aldosterone concentrations relative to Ang II (AA2-R) over gestation in NT (P<0.010) but not HYP patients (p=0.141, Table 2).

**Figure 2.**
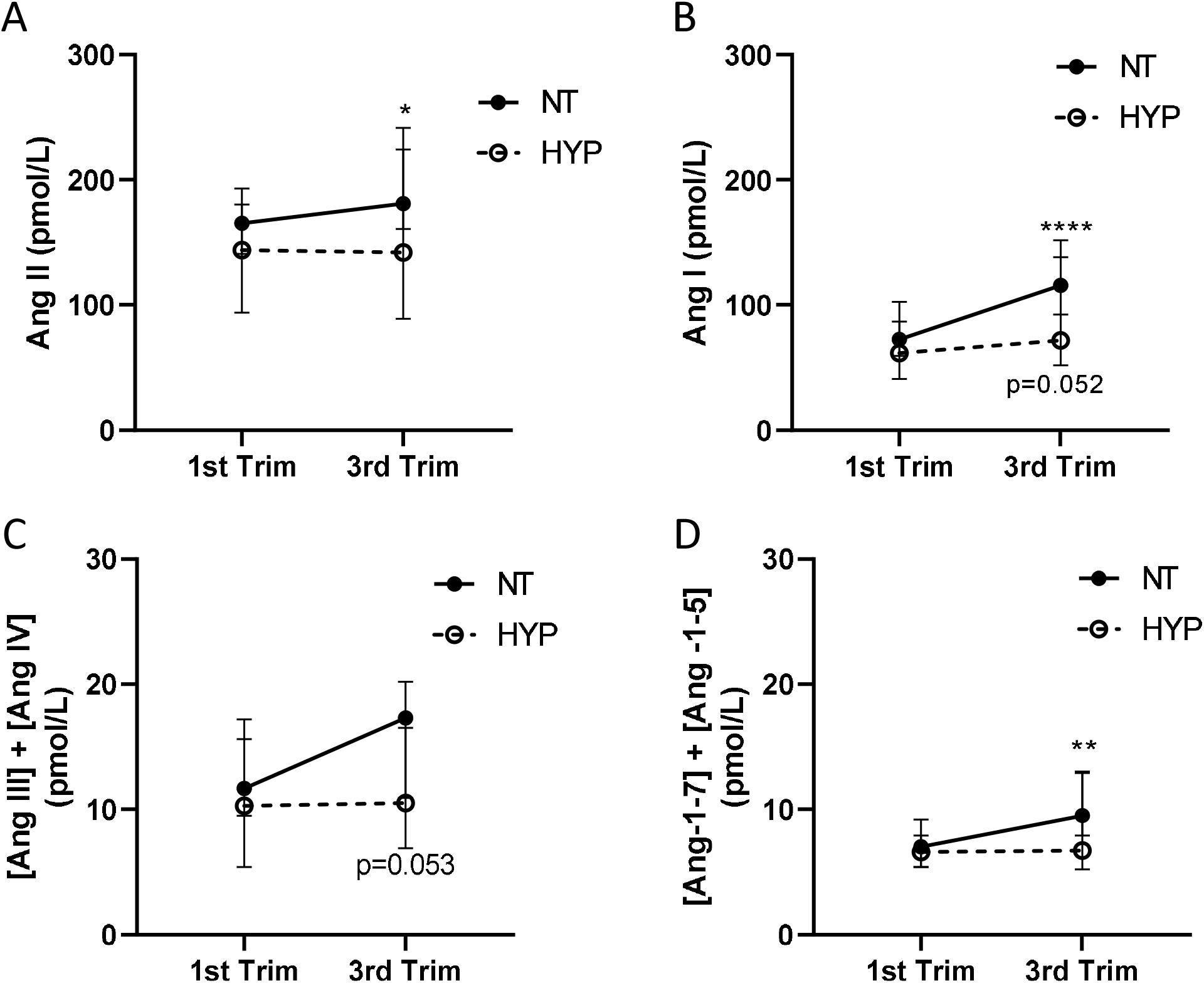
Equilibrium concentrations of Ang II, and upstream and downstream angiotensin metabolites in normotensive and hypertensive pregnancy. A) Ang II and B) Ang I concentrations in the first and third trimester in NT (n=47) and HYP (n=27) groups. Data are median with 95^th^ confidence intervals (CI). *, P<0.01, ****, P<0.01, within-group increase from first to third trimester analyzed by Wilcoxon matched-pairs signed rank tests. P=0.052 increase across gestation in HYP. C) The concentration of n-terminal metabolites of Ang II, calculated from the sum of the concentrations of Ang III and Ang IV, and D) the concentration of c-terminal metabolites of Ang II, calculated from the sum of the concentrations of Ang-1-5 and Ang-1-7. Data are median with 95^th^ CI. P=0.053, between group analysis at the third trimester analyzed via Mann-Whitney. **, P<0.01 within-group increase from first to third trimester analyzed by Wilcoxon matched-pairs signed rank tests.

Metabolism of Ang II was assessed by examining the concentrations of n-(sum of the concentrations of Ang III and Ang IV) and c-terminal metabolites (sum of the concentrations of Ang-1-5 and Ang-1-7), where c-terminal metabolites are reflective of activity of the counter-regulatory arm of the RAAS[25]. Concentrations in pmol/L of n- and c-terminal metabolites of Ang II were 10-fold lower than Ang II and Ang I concentrations in both NT and HYP groups (Figure 2B, 2C, Table 2). Concentrations of Ang-1-7 were below the LLOQ in most patients in the NT group (detected in only 7 and 16 patients in the first and third trimester, respectively), and in the HYP group (detected in only 7 and 9 patients in the first and third trimester, respectively), so median values with IQR for this analyte were less than the LLOQ and not listed in Table 2. Half the value of the LLOQ (a concentration of 1.5 pmol/L) in those samples with Ang-1-7 concentrations below the LLOQ was used to calculate the sum of c-terminal peptides. The concentrations of c- and n-terminal metabolites were not different in NT versus HYP in the first or third trimester (Figure 2C, 2D), but in the NT group, the concentration of c-terminal metabolites significantly increased over gestation (Figure 2D).

## 4. Discussion

Previous studies have demonstrated that the RAAS is suppressed in pregnancies with pregnancy hypertension, but deviations in the activity of the RAAS compared to normotensive pregnancy have not been comprehensively described. We used RAS Fingerprint to quantify multiple angiotensin metabolites and biomarkers of the RAAS over gestation in pregnant women with or without pregnancy hypertension. The main results from this study, the first to report LC-MS/MS-based RAAS profiles in a prospective study of pregnancy hypertension, are 1) a marked increase across gestation was evident in PRA-S, and especially aldosterone concentrations, in NT pregnancy, but 2) not in those that developed HYP, where PRA-S was largely unchanged after the first trimester of pregnancy, and third trimester aldosterone concentrations were significantly lower than the NT group; 3) the increase in aldosterone secretion across gestation was proportionally greater than the change in PRA-S or Ang II in both groups, with a more robust effect in the NT group; and 4) c- and n-terminal metabolites, present in low concentrations in both groups, generally increased over gestation in the NT but the HYP group. Taken together, these findings indicate that normotensive pregnancy is associated with a sustained increased in Ang I (and to a lesser degree, Ang II) concentrations over gestation, accompanied by a marked elevation in aldosterone concentrations and moderate increase in alternative RAAS metabolites. The predominate features of pregnancies that develop HYP are stalled or waning activation of the RAAS in the second half of pregnancy (accompanied by unchanging levels of angiotensin peptides) and attenuated secretion of aldosterone.

The comprehensive RAAS profiles generated in our study agree with previous studies reporting low renin and aldosterone levels in normal and hypertensive pregnancy, and extend them to include detailed activity of angiotensin peptide metabolites over gestation. PRA and aldosterone concentrations are progressively increased with pregnancy, and can exceed non-pregnant levels by 2-3 fold[26, 52]. PRA is increased early in gestation [53], where aldosterone concentrations peak in the third trimester[54]. In pregnancies that develop hypertension, some studies have shown that PRA is initially elevated as in normotensive pregnancy, but fails to increase over gestation or decreases to non-pregnant levels[25]. Several studies have demonstrated that, aldosterone concentrations, although reduced compared to normotensive, still rise to some degree over pregnancy, even when PRA is declining[25, 26]. These studies are consistent with our observations, where PRA-S and aldosterone increased over gestation only in the NT group, but aldosterone remained elevated in in the HYP group. Our study did not contain a non-pregnant group for comparison, but reported values of RAAS biomarkers in non-pregnant cohorts generated with the same LC-MS/MS-based methodology are available. The median and IQR reported for PRA-S and Ang II in a cohort of 77 individuals with hypertension was 165.6 (80.6–328.3) pmol/L and 100.9 (56.3–227.0) pmol/L, respectively[32]. These values are substantially lower than PRA-S and Ang II values reported here in the third trimester of the HYP group, suggesting that RAAS activity was still higher compared to non-pregnant state.

Because pre-eclampsia is associated with lower plasma volume [54], suppression of the RAAS is somewhat paradoxical. In longitudinal studies of pregnancies that developed gestational hypertension or pre-eclampsia, downward trending of PRA levels over pregnancy correlated with the onset of high blood pressure [25, 26, 55]. In our study, patients that developed HYP failed to increase RAS components over pregnancy, and had generally lower concentrations of angiotensin peptides in the third trimester. Downregulation of renin activity with disease course suggests a compensatory response to high blood pressure[55], rather than a primary event in the pathogenesis of the disease. An additional explanation for low activation of the RAAS in hypertensive pregnancies could be reduced concentration or availability of AGT[56], the substrate for renin. Langer et al previously made the observation that first trimester Ang I levels were low compared to Ang II (similar to our study), and suggested that Ang II synthesis was not limited by renin, but rather perhaps by AGT [52].

Historical studies by Gant et al demonstrate that normotensive pregnancies become refractory to the pressor effects of Ang II, presumably for protection against vasoconstrictive effects of Ang II, where pregnancies that develop hypertension progressively lose this response over the second half of pregnancy [57]. This suggests factors impacting AT_1_ receptor signaling in pregnancy, such as presence or absence of AT_2_ receptors[58], metabolic derangements{34658252}, or AT_1_ autoantibody-binding might facilitate negative cellular effects of Ang II (promotion of inflammation[59] or renal[60]), that lead to the progression of hypertension. In our cohort, blood pressure was elevated in HYP compared to NT in the first trimester, suggesting the presence of underlying vascular dysfunction in this group, potentially caused by previous pre-eclampsia or type 1 diabetes exhibited in this group.

Factors contributing to aldosterone secretion in hypertensive pregnancy are complex. Previous studies reported that aldosterone concentrations in pregnancy exceeded those expected on the basis of renin levels[57]. In our study, aldosterone concentrations relative to Ang II were significantly increased over gestation only in NT groups, but HYP pregnancies still had elevated aldosterone compared to non-pregnant cohorts[32] in the second half of pregnancy, despite no sustained activation of the RAAS. These data suggest non-Ang II stimulation of aldosterone is a feature of both normotensive and hypertensive pregnancy, but the possibility of altered sensitivity of AT_1_ receptors to Ang II in the adrenal cortex in one group compared to another is not excluded. Purported non-angiotensin stimulants of aldosterone in pregnancy include ACTH, where ACTH simulated aldosterone not only in normotensive but also hypertensive pregnancies[61], and VEGF [62]. In pre-eclampsia, the presence of circulating factors such as the VEGF receptor binding splice variant, sFlts-1, and AT_1_-autoantibodies, may impair aldosterone synthesis or release[63]. In addition, aldosterone concentrations could be suppressed by excess sodium in pre-eclampsia resulting from activation of epithelial sodium channels (ENaC)[64]. Notably, gain of function variants in aldosterone synthase (CYP11B2) were preferably present in normotensive versus pre-eclamptic pregnancies, suggesting an evolutionary basis for an active mineralocorticoid receptor system to protect against pre-eclampsia[65].

Enhanced concentrations of Ang-1-7 were reported in normal pregnancy, and reduced concentrations in pre-eclampsia, where authors hypothesized that Ang-1-7 may function in healthy pregnancy to oppose potentially adverse (vasoconstrictive) effects of Ang II[29, 66]. Similarly, in a recent study, plasma Ang-(1-7) concentrations were elevated throughout pregnancy compared to a non-pregnant control group, and reduced in preeclampsia[28], suggesting a protective effect of ACE2 against hypertension during pregnancy via generation of Ang-1-7. In contrast, we report limited detection of Ang-1-7 in NT and HYP patients (using mass spectrometry, which is the gold standard for quantification of biological molecules and a major strength of our study[67, 68]); in samples where Ang-1-7 was detected, concentrations were significantly lower than those reported in other studies via radioimmunoassay[28, 29]. Discrepancies in reported values likely result from different methodologies used for the detection of Ang-1-7, where cross-reactivity in antibody-based assays, (as peptide sequences may only differ by one or two amino acids), may explain artificially high measured concentrations of Ang-1-7[69]. Notably, reliability of ELISA for quantification of angiotensin-(1-7) has recently been called into question[70]). Ang-1-5, a c-terminal metabolite of Ang-1-7, is considered a component of the alternative RAAS[51, 71]. Ang-1-5 was readily detected in our study by mass spectrometry, so we used the sum of Ang-1-7 and Ang-1-6 concentrations as an indicator of the alternative RAAS. Concentration of Ang-1-7 + Ang-1-5 did increase over gestation in NT, but not HYP pregnancies. However, concentrations of angiotensin metabolites downstream from Ang II, (whether c- or n-terminally degraded), 10-fold less than the concentrations of Ang II and Ang I. This data suggests the main bioactive effector of the RAAS is Ang II, and that modulatory effects of Ang-1-7 in pregnancy are likely specific to local tissues[72].

Our study has several limitations. We did not differentiate among pre-eclampsia and gestational hypertension due to small numbers. Pre-eclampsia, especially, is a heterogenous disease, where some subtypes may result from a different pathophysiology[6], with a different RAAS profile. Also, our study did not control for sodium or volume status, which could affect concentrations of aldosterone and PRA-S. Although previous studies have indicated that sensitivity of renin and aldosterone to salt are blunted in pregnancy[57, 73], differences in salt and aldosterone relationships in pregnancy hypertension versus normotensive are not well-understood.

## 5. Conclusions

In summary, we sought to define prominent features of RAAS activity associated with the development of pregnancy hypertension using a novel methodology for comprehensive assessment of the RAAS. Concentrations of PRA-S, angiotensin peptides, and aldosterone were not different among NT and HYP in the first trimester, significantly increased over gestation only in the group that remained NT, and tended to be lower in the third trimester in HYP compared to NT. We conclude that pregnancies that develop HYP are characterized by stalled or waning activation of the RAAS in the second half of pregnancy (accompanied by unchanging levels of angiotensin peptides) and attenuated secretion of aldosterone. Clinical application of this knowledge may inform detection or treatment of pregnancy hypertension or related adverse outcomes.

## Author contributions

Study conception and design (JAB, JOB, AS); clinical data collection (KV, AS, HH, CC); blood analysis of RAS and interpretation (RS, MP); analysis and interpretation of results (RS, MP, AS, JAB, JOB); manuscript preparation (RS).

## Institutional Review Board Statement

The study was conducted in accordance with the Declaration of Helsinki, and approved by the Institutional Review Board at the University of Kentucky. All study procedures/methods were performed in accordance with relevant guidelines and regulations.

## Informed Consent Statement

Informed consent was obtained from all subjects involved in the study.

## Data availability

Data may be made available by investigators upon reasonable request to corresponding author.

## Data Availability

All data produced in the present study are available upon reasonable request to the authors

